# MODERN CONTRACEPTIVE USE AMONG WOMEN IN ZAMBIA: A DESCRIPTIVE SURVEY

**DOI:** 10.1101/2021.07.13.21259719

**Authors:** Chilambwe Jully Mwansa, Tyson Chipokoso, Melvin Mwansa, Mowa Zambwe

## Abstract

**OBJECTIVE:** This study aimed to describe the modern contraceptive use by women of childbearing age in Zambia.

**METHODS:** The study was retrospective descriptive observational design. Secondary data of 13,683 women aged 15 - 49 who participated in the ZDHS 2018 were analysed. Data were extracted using a data extraction tool and analysed using IBM SPSS Statistics 26. Since the ZDHS 2018 used numeric data, the study was quantitative research. The study examined the association between demographic and socioeconomic characteristics and modern contraceptive use using Pearson, Chi-Square and multiple logistic regression.

**RESULTS:** In Zambia, the most used type of modern contraception were injectables (52%). Female condom is the most known type of modern contraceptives (55%). The significant factors associated with the use of modern contraceptives were age group 25-29 and 45-49 (AOR=1.41 95% CI=1.05, 1.90) and (AOR=0.41, 95% CI 0.28, 0.62), respectively. Being married and widowed (AOR=2.18, 95% CI 1.75, 2.71), and (AOR=0.50, 95% CI 0.31, 0.78), respectively. The likelihood utilization varied across the gradient of parity, educational attainment, and wealth. Educated and wealthy women were more likely to use modern methods compared to women with no education and in the poor category, respectively.

**CONCLUSION:** The study established that factors such as age, marital status, the number of living children (parity), religion, and education level and wealth index remain significant issues in determining modern contraceptive use among childbearing women aged 15 to 49 in Zambia. Therefore, concerted efforts are required to increase use of modern methods of contraception by addressing these determinants.

**ARTICLE SUMMARY:** *Strengths and limitations of the study:*

1. The study utilized secondary data from the ZDHS 2018 as a proxy to establish the determinants of contraceptive use among women of childbearing age 15-49 in Zambia and therefore added to the body of knowledge.
2. This study was limited by its cross-sectional nature and hence causal inferences cannot be made.
3. The study relied on self-report measures, which could be affected by social desirability bias or memory bias

## INTRODUCTION

Modern contraceptive use remains an important public health intervention, a cost-effective strategy to reduce maternal mortality and avert unintended pregnancies, especially in developing countries. Despite these benefits, there are reports of low usage among reproductive-aged women in most developing countries.^1^ The United Nations Population Fund projects a mean global population growth of 9.7 billion and 10.9 billion in 2050 and 2100 respectively.^2^ Among the regions and countries whose populations are expected to contribute to this upsurge, Sub-Saharan Africa is projected to contribute to more than half of that.^2^

UNFPA has recorded a TFR of 2.5 and 4.6 for global and Sub-Saharan Africa, respectively. The level of total fertility for a given period reflects the average number of children women would bear if current fertility rates remained unchanged during their reproductive lifespans (15-49 years of age).^3^ Zambia’s TFR stands at 4.7; higher in the rural of 5.8 and lower in the urban of 3.4.^4^

Fertility is central to population health and is defined as, the natural ability to conceive offspring.^5^ High fertility, however, has effects that will adversely impede the achievement of the SDGs of eradicating hunger and poverty, achieving gender equality, and improving health and education.^2^ This, therefore, calls for concerted efforts to control population growth through FP

Family Planning is among the indicators of the SDGs, Indicator 3.7.1 which stipulates, the proportion of women of reproductive age (aged 15-49 years) who have their need for family planning satisfied with modern contraceptives by 2030.^6^

Zambia, being one of the original FP 2020 countries, set a goal to increase the mCPR to 58 percent by 2020.^7^ Furthermore, the country has an active FP Costed Implementation Plan and several proactive initiatives to increase the mCPR, however Zambia’s unmet need for contraception for all women remains at 22 percent despite substantial increases in mCPR from 33 percent in 2007 to 48.5 percent in 2020.^8,9^

Contraception describes pregnancy prevention by inhibiting the normal process of ovulation, fertilisation, and implantation. Varied modern contraceptive methods have been developed including male and female condoms, oral hormonal pills, IUD, implants, male and female sterilization (vasectomy and tubal ligation), injectables, vaginal barriers, and emergency contraception. The use of modern contraceptive methods allows couples and individuals to attain their desired number of children and determine the spacing of pregnancies. This helps to reduce maternal deaths and child mortality by preventing unsafe abortions, birth injuries, and all other complications that happen due to pregnancy.^10^

Based on the world contraceptive use by method (any, modern or traditional) of 2019 for 1994 and 2019, *a*mong the 1.9 billion women of reproductive age (15-49 years), 1.1 billion need family planning. 842 million use modern methods of contraception. The proportion of women who need family planning satisfied by modern methods (Sustainable Development Goals indicator 3.7.1) is 76 percent in 2019.^11^

## MATERIALS AND METHODS

The study was descriptive and adopted a retrospective observational study design. It extracted data for women of childbearing age, 15-49 from the 2018 Zambia Demographic and Health Survey database. The dataset provided data on the demographic and socioeconomic status such as age, parity, marital status, place of residence, education attainment, religion wealth index knowledge and attitude on modern contraceptive use among women of childbearing age 15-49 in Zambia.

### Sampling and data collection methods

The sampling frame used for the 2018 ZDHS was the Census of Population and Housing (CPH) of the Republic of Zambia, conducted in 2010 by ZamStats. Zambia was divided into 10 provinces. Each province was subdivided into districts, each district into constituencies, and each constituency into wards. In addition to these administrative units, during the 2010 CPH, each ward was divided into convenient areas called census supervisory areas (CSAs), and in turn, each CSA was divided into enumeration areas (EAs). According to the Zambian census frame, each EA consists of an average of 110 households.

### Statistical analysis

The analysis of this study used secondary data (ZDHS 2018 Dataset). IBM SPSS statistics 26 was used for data analysis. Frequency distribution tables and graphs such as bar graphs and pie charts were used to display results. The dataset for women age 15-49 (ZMIR71FL) was used and applied sample weights to adjust the sample.

Analysis of the data was done at three stages namely, Univariate, Bivariate (Chi-square), and using logistic regression analysis. Descriptive analysis at the univariate stage was performed. Frequencies of the different categories of the background characteristics of respondents were presented.

The inferential analysis was done after cross-tabulation was performed to establish the association between the independent variables (socio-economic and demographic factors) with the dependent variable (Modern contraceptive use). Statistical significance test (chi-square) was performed to estimate whether the results observed in the analysis were by chance or of statistical significance which was done by using the hypothesis tests.

Binary logistic regression was used to determine the likelihood of using modern contraceptives across all the factors (demographic and socioeconomic factors). The dependent variable (modern contraceptive use) was treated as dichotomous for it had only two outcomes either no or yes coded as 0 or 1, respectively. In this case, a woman could either use modern contraceptives or not.

## RESULTS

The 2018 ZDHS collected information about contraception. This paper presents the findings concerning modern contraceptive use among women of childbearing age 15-49. It includes all women who used modern contraceptives from rural and urban areas in Zambia.

### Background characteristics of respondents

Table 1 below shows the background characteristics of respondents in the ZDHS 2018. Respondents in the age group 15-19 had the highest proportion (21 percent) of women selected and interviewed in the survey. The age group 45-49 had the lowest proportion (7 percent) of respondents. By marital status, more than half of the married women (56 percent) were interviewed. The least proportion of respondents (1 percent) were cohabiting.

**Table 1:** Background characteristics of respondents.

*Percent distribution of women age 15 to 49 by selected background characteristic, Zambia ZDHS 2018*
| Background characteristics | Freq. | Percent |
| --- | --- | --- |
| **Age** |  |  |
| 15-19 | 2,036 | 21.4 |
| 20-24 | 1,863 | 19.6 |
| 25-29 | 1,611 | 16.9 |
| 30-34 | 1,309 | 13.8 |
| 35-39 | 1,178 | 12.4 |
| 40-44 | 867 | 9.1 |
| 45-49 | 640 | 6.7 |
| **Marital status** |  |  |
| Never married | 2,905 | 30.6 |
| Married | 5,338 | 56.2 |
| Living together | 46 | 0.5 |
| Widowed | 267 | 2.8 |
| Divorced | 676 | 7.1 |
| Separated | 271 | 2.8 |
| **Children Ever Born** |  |  |
| No child | 2,311 | 24.3 |
| 1-4 | 4,899 | 51.6 |
| 5-9 | 2,107 | 22.2 |
| 10-14 | 185 | 2.0 |
| **Religion** |  |  |
| Catholic | 1,642 | 17.3 |
| Protestant | 7,688 | 80.9 |
| Muslim | 45 | 0.5 |
| Other | 128 | 1.4 |
| **Residence** |  |  |
| Urban | 4,383 | 46.1 |
| Rural | 5,120 | 53.9 |
| **Region** |  |  |
| Central | 821 | 8.6 |
| Copperbelt | 1,481 | 15.6 |
| Eastern | 1,095 | 11.5 |
| Luapula | 745 | 7.8 |
| Lusaka | 1,931 | 20.3 |
| Muchinga | 535 | 5.6 |
| Northern | 727 | 7.7 |
| North western | 521 | 5.5 |
| Southern | 1,105 | 11.6 |
| Western | 541 | 5.7 |
| **Education attainment** |  |  |
| No education | 754 | 7.9 |
| Primary | 4,161 | 43.8 |
| Secondary | 4,052 | 42.6 |
| Higher | 536 | 5.6 |
| **Wealth index** |  |  |
| poor | 3,371 | 35.5 |
| Middle | 1,751 | 18.4 |
| Rich | 4,381 | 46.1 |
| **Total** | **9,503** | **100.0** |

Slightly more than half of the respondents had 1 to 4 children (52 percent), while only 2 percent of the respondents had 10 to 14 children. Most of the women were Protestant with 81 percent distribution and the lowest were Muslim women with 1 percent distribution. By place of residence, more than half of the women aged 15-49 (54 percent) were from rural areas, while only 46 percent were from urban areas.

The majority of women by province were from Lusaka province (20 percent) followed by Copperbelt province (16 percent) and the least was from Muchinga province (6 percent) and Western province (6 percent). The highest proportion (44 percent) of the respondents had attained primary education, and the lowest were those who had higher education at 6 percent. The majority of the respondents (46 percent) were those from rich households, and the lowest were those from the middle wealth index at 18 percent.

### Type of modern contraceptives used among women of childbearing age 15-49

Figure 2 below shows the type of modern contraceptives used by women aged 15 to 49 in Zambia. Slightly more than half of the women aged 15 to 49 (52 percent) used injectables, and the least used male sterilization and other modern methods at 0.1 percent, respectively.

**Figure 1:**
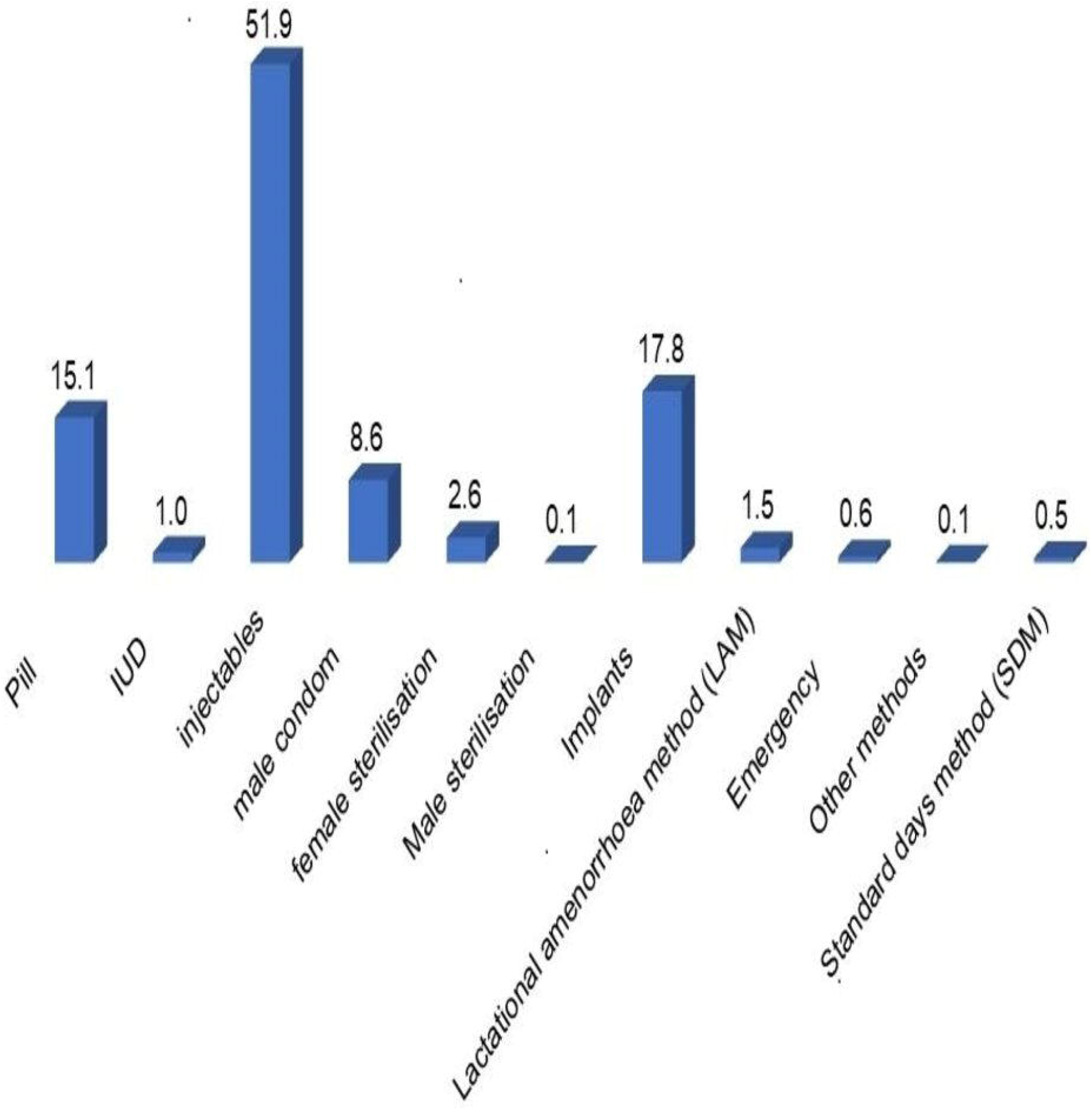
Profile of contraceptive methods used among women by percentage.

**Figure 2:**
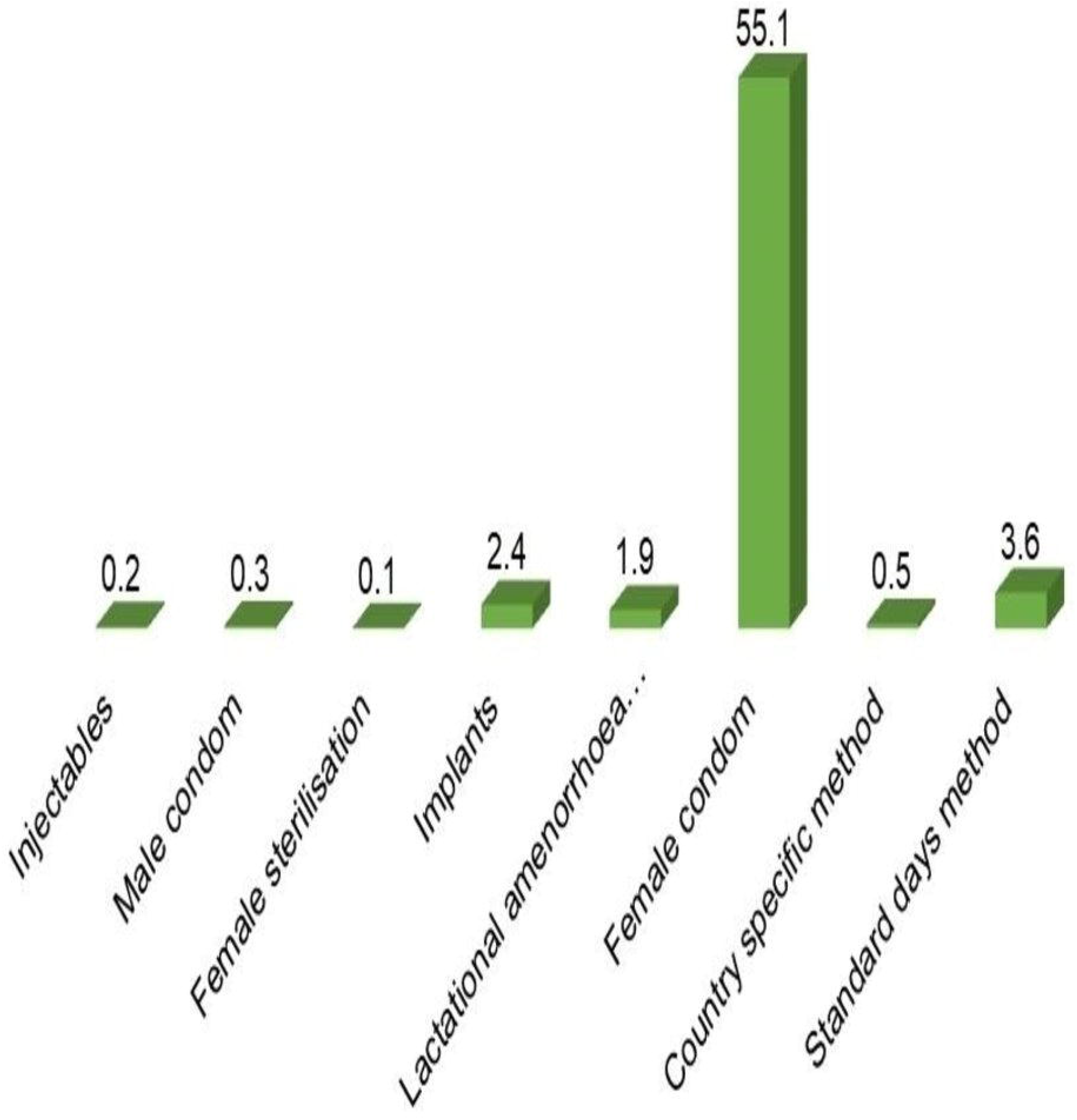
Knowledge and attitude on modern contraceptive use among women aged 15-49 by percentage.

**Figure 3:**
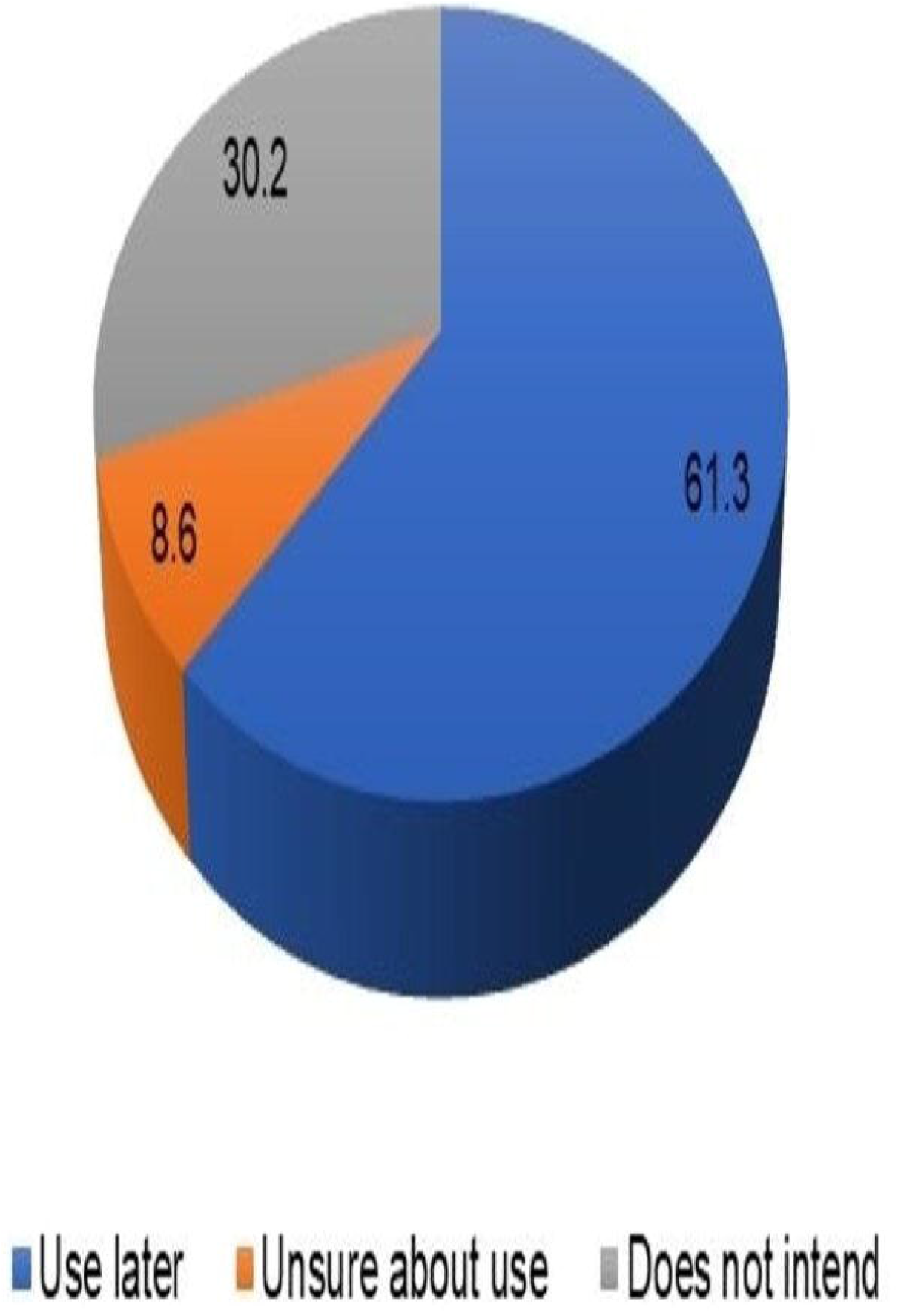
Attitudes of Women aged 15 to 49 attitudes towards the use of modern contraceptives.

### Effect of Demographic and socioeconomic status of women on modern contraceptive use

Table 3 below shows the demographic and socioeconomic characteristics of women aged 15-49 in the ZDHS 2018 who used modern contraceptives and the determinants thereof. Use of modern contraceptives among women aged 15-49 increases with increasing age in the young ages (15 to 34) and reduces in the old ages (35 to 49). Most women aged 15 to 49 who used modern contraceptives were from the age group 30-34 (48 percent), and the least were from the age group 15-19 at 13 percent. Further, the P-value (<0.05), indicates that there is a significant relationship between women’s age and use of modern contraceptives. Therefore, there is enough evidence to conclude that women’s age influences the use of modern contraceptives in Zambia.

**Table 2:** Modern contraceptive use among women aged 15 to 49 by background characteristics.

| Demographic and socioeconomic characteristics | Percent distribution of women who used modern contraceptives |  | P-value |
| --- | --- | --- | --- |
|  | Did not use modern contraceptives | used modern contraceptives |  |
| **Age** |  |  |  |
| 15-19 | 1782 (87.5) | 254 (12.5) | **0.000** |
| 20-24 | 114 (64.1) | 668 (35.9) |  |
| 25-29 | 868 (53.) | 742 (46.1) |  |
| 30-34 | 686 (52.4) | 624 (47.6) |  |
| 35-39 | 635 (53.9) | 543 (46.1) |  |
| 40-44 | 528 (61.0) | 338 (39.0) |  |
| 45-49 | 477 (74.5) | 163 (25.5) |  |
| **Marital status** |  |  |  |
| Never married | 2516 (86.6) | 389 (13.4) | **0.000** |
| Married | 2757 (51.6) | 2581 (48.4) |  |
| Cohabiting | 28 (62) | 17 (3.7) |  |
| Widowed | 228 (85) | 40 (14.8) |  |
| Divorced | 465 (68.7) | 212 (31.3) |  |
| Separated | 177 (65.4) | 93 (34.6) |  |
| **Parity** |  |  |  |
| No child | 2168 (3.8) | 143 (6.17) | **0.000** |
| 1-4 | 2734 (55.8) | 2165 (44.2) |  |
| 5-9 | 1155 (54.) | 952 (45.2) |  |
| 10-14 | 114 (61.4) | 72 (38.6) |  |
| **Religion** |  |  |  |
| Catholic | 1115 (67.) | 527 (32.1) | **0.019** |
| Protestant | 4943 (64.3) | 2745 (35.7) |  |
| Muslim | 22 (48.9) | 23 (51.1) |  |
| Other | 91 (70.7) | 38 (29.3) |  |
| **Residence** |  |  |  |
| Urban | 2826 (64.5) | 1557 (35.5) | **0.532** |
| Rural | 3345 (65.3) | 1775 (34.7) |  |
| **Educational attainment** |  |  |  |
| No education | 523 (69.3) | 232 (30.9) | **0.056** |
| Primary | 2642 (63.5) | 1518 (36.5) |  |
| Secondary | 2657 (65.6) | 1395 (34.4) |  |
| Higher | 349 (65.1) | 187 (34.9) |  |
| **Wealth index** |  |  |  |
| Poor | 2246 (66.6) | 1125 (33.4) | **0.015** |
| Middle | 1072 (61.2) | 679 (38.8) |  |
| Rich | 2853 (65.1) | 1529 (34.9) |  |

*Source: ZDHS 2018*

**Table 3:** the odds of demographic and socioeconomic factors and the use of modern contraceptives.

| Characteristics | AOR | 95% CI |  |
| --- | --- | --- | --- |
| **Age** |  |  |  |
| 15-19 | 1 |  |  |
| 20-24 | 1.28 | (0.98 | 1.67) |
| 25-29 | 1.41** | (1.05 | 1.90) |
| 30-34 | 1.24 | (0.91 | 1.69) |
| 35-39 | 0.98 | (0.69 | 1.39) |
| 40-44 | 0.77 | (0.55 | 1.07) |
| 45-49 | 0.41** | (0.28 | 0.62) |
| **Parity** |  |  |  |
| No child | 1 |  |  |
| 1-4 | 7.89** | (5.01 | 12.42) |
| 5-9 | 11.91** | (7.04 | 20.15) |
| 10-14 | 14.51** | (7.68 | 27.43) |
| **Marital status** |  |  |  |
| Never married | 1 |  |  |
| Married | 2.18** | (1.75 | 2.71) |
| Cohabiting | 1.4 | (0.55 | 3.56) |
| Widowed | 0.5** | (0.31 | 0.78) |
| Divorced | 1.08 | (0.80 | 1.47) |
| Separated | 1.15 | (0.73 | 1.79) |
| **Religion** |  |  |  |
| Catholic | 1 |  |  |
| Protestant | 1.07 | (0.91 | 1.27) |
| Muslim | 1.82 | (0.88 | 3.81) |
| Other | 0.79 | (0.48 | 1.29) |
| **Residence** |  |  |  |
| Urban | 1 |  |  |
| Rural | 0.89 | (0.72 | 1.08) |
| **Educational attainment** |  |  |  |
| No education | 1 |  |  |
| Primary | 1.46** | (1.19 | 1.79) |
| Secondary | 1.78** | (1.14 | 1.63) |
| Higher | 1.71** | (1.05 | 1.57) |
| **Wealth index** |  |  |  |
| Poor | 1 |  |  |
| Middle | 1.36** | (1.14 | 1.63) |
| Rich | 1.28** | (1.05 | 1.57) |

Source: ZDHS 2018
Significant at <0.05\*\*

By marital status, most of the women aged 15-49 who used modern contraceptives were married (48 percent), and the minority were cohabiting (4 percent). Further, the P-value (<0.05), indicates that there is a significant relationship between women’s marital status and the use of modern contraceptives. Therefore, there is enough evidence to conclude that women’s marital status influences the use of modern contraceptives in Zambia.

Many of the women aged 15 to 49 who used modern contraceptives had parity (5-9 children), and the lowest use of modern contraceptives was with women who had no child (6 percent). Further, the P-value (<0.05), indicates that there is a statistically significant relationship between women’s parity status and the use of modern contraceptives. Therefore, there is enough evidence to conclude that women’s parity influences the use of modern contraceptives in Zambia.

Slightly more than half (51 percent) of women aged 15 to 49 who used modern contraceptives were Muslim and the least belonged to other religions (29 percent). Further, the P-value (<0.05), indicates that there is a statistically significant association between women’s religious affiliation and use of modern contraceptives. Therefore, there is enough evidence to conclude that women’s religious affiliation influences the use of modern contraceptives in Zambia.

By place of residence, there is a slight difference in the use of modern contraceptives between women who reside in urban areas and rural areas at 36 percent, and 35 percent, respectively. However, the observed P-value (>0.05), indicates that there is no statistically significant association between the place of residence of women and the use of modern contraceptives. There is enough evidence to conclude that place of residence does not influence the use of modern contraceptives in Zambia.

Women who had attained primary education had the highest use of modern contraceptives (37 percent), while the lowest use was among women with no education (30 percent). The observed P-value (<0.05), indicated that there was a statistically significant association between educational attainment and the use of modern contraceptives. Therefore, there is enough evidence to conclude that educational attainment does influence the use of modern contraceptives in Zambia.

Further, women from middle-income households had used modern contraceptives the most (38 percent) and those from poor households (33 percent) used the least modern contraceptives. The P-value (<0.05) observed indicated that there was a statistically significant relationship between the wealth status of women aged 15-49 and the use of modern contraceptives in Zambia. Therefore, there is enough evidence to conclude that the wealth status of women aged 15 to 49 influences the use of modern contraceptives.

### Knowledge and attitude on modern contraceptive use among women aged 15-49

Figure 5 below shows women aged 15 to 49 who knew any type of modern contraceptives. More than half (55 percent) of the women reported having known female condoms as the type of modern contraceptive, while only 0.1 percent know about female sterilization.

### Women aged 15 to 49 attitudes towards the use of modern contraceptives

Figure 6 below shows women aged 15 to 49 attitudes towards the use of modern contraceptives in Zambia. The majority of the women intended to use modern contraceptives later (61 percent), while the minority were unsure about using modern contraceptives (9 percent).

### Binary multiple logistic regression between background characteristics and modern contraceptive use

Among women who were in the age group 25-29, the odds of using modern contraceptives were 41 percent higher, compared to those in the age group 15-19 (AOR 1.41, 95% CI 1.05, 1.90). Further, for women in the age group 45-49, the odds of using modern contraceptives were 59 percent lower than those in the age group 15-19 (AOR 0.41, 95% CI 0.28, 0.62). In terms of parity, the odds of using modern contraceptives increased as the number of children ever born per woman increased. Among women who had children between10-14 were 15 times more likely to use modern contraceptives compared to those who had no child (AOR 14.51, 95% CI 7.68, 27.43).

By marital status, women who were married were 2 times more likely to use modern contraceptives compared to never-married (AOR 2.18, 95% CI 1.75, 2.71). Among women who were widowed, the odds of using modern contraceptives were 50 percent lower, compared to those who were never married (AOR 0.50, 95% CI 0.31, 0.78). There was no statistically significant relationship between women’s religious affiliation and the use of modern contraceptives. Further, the association between place of residence and the use of modern contraceptives was not statistically significant.

Among women who had attained secondary education, the odds of using modern contraceptives were 78 percent higher, compared to those who had no education (AOR 1.78, 95% CI 1.14, 1.63). Further, among women who were in middle-income households, the odds of using modern contraceptives were 36 percent higher, compared to those from poor households (AOR 1.36, 95% CI 1.14, 1.63). For women who came from rich households, the odds of using modern contraceptives were 28 percent higher than those who came from poor households (AOR 1.28, 95% CI 1.05, 1.57).

## DISCUSSION

The main objective of this study was to explore the use of modern contraceptives among women of childbearing age (15-49) in Zambia during the period 2018-2019. Specifically, the study examined the prevalence of modern contraceptive methods used by Zambian women and the factors that might influence their use. It also identified demographic and socioeconomic factors that influenced modern contraceptive use among Zambian women.

Among women who were in the age group 25-29, the odds of using modern contraceptives were 41 percent higher, compared to those in the age group 15-19 (AOR 1.41, 95% CI 1.05, 1.90). Further, for women in the age group 45-49, the odds of using modern contraceptives were 59 percent lower than those in the age group 15-19 (AOR 0.41, 95% CI 0.28, 0.62). There was a significant relationship between age, particularly, older women, and the use of modern contraceptives. Similarly, a study conducted in Indonesia revealed that women’s age, especially older age, was significantly associated with contraceptive use among married women.^12^ This result was consistent with the other studies conducted in Ghana and Nigeria, documenting that older woman had a lower level of concern with modern contraceptive use. This finding was related to their lower fecundity rates and less active sexual desires. The benefits of using contraception were to delay or space subsequent pregnancies and to limit the number of children. Although older women used contraception less often than young women, they still considered the use of contraceptives; of note, only a small proportion of women aged 35–49 reported having gone through menopause.^13^

This study shows that there is a direct significant association between parity and the use of modern contraceptives. It is observed that as the number of children ever born increases, the utilization of modern contraceptives increases too. This is consistent with a study conducted in Indonesia, Women with five or more living children were more likely to use contraceptives.^12^ This finding is consistent with those studies performed in Ghana. The addition of one child would increase the tendency of married women to use contraception.^13^

Further, the findings of this study showed that there was no association between modern contraceptive use and the religious affiliation of women in Zambia. However, a study conducted in Nigeria revealed that Catholic women, other Christians, Traditionalists, and respondents from other denominations were significantly more likely to use modern contraceptives compared to Muslim women. This is so because it has been shown that Muslims have a lower level of education as compared to Christians, as such the latter are more subjected to information about the importance of utilization of FP services.^14^ Another study conducted in Ghana, indicated that women affiliated to the Muslim faith were also more likely to use contraceptive method compared to women of no religion. This is also interesting because the faith seems to admonish its followers to procreate and abound in number.^15^ However, the Muslim faith does not prohibit FP; in fact, many Islamic scholars approve of it especially where the well-being of the family may be compromised (ibid).

There was no significant relationship between modern contraceptive use and place of residence in Zambia. This is in contrast with the study conducted in Nigeria, in which women residing in urban areas were significantly more likely to currently use modern contraceptive methods compared to the women who lived in rural areas (AOR=1.223, 95% CI 1.111, 1.346). A possible explanation could be, as women who lived in urban areas had a greater chance of being more educated, have more access to health facilities, greater exposure to mass media messages and more knowledge about modern contraceptive issues than their counterpart who dwelt in the rural areas where unmet need for FP was high.^14^

The findings also showed that education was an important variable for modern contraceptive usage. There was a direct relationship between modern contraceptive use and levels of education of the women as observed that women who had higher, secondary and primary education were all more likely to use modern contraceptive than their counterparts who had no form of education in Zambia. The finding agrees with other findings.^16^ argued that a certain educational threshold is needed to activate appreciable use of contraceptives. According to the study conducted in Ghana, female education did not only improve women’s status but was also important for achieving their reproductive health needs. Thus, women who had higher education were three times likely to be contraceptive users compared to their counterparts who had no education (ibid).

Similar patterns have been shown in studies conducted in Bangladesh and Ghana that, education had an extremely significant influence on contraceptive use. Women with higher education were more likely to use contraception than those without formal education.^17^ This finding was the result of highly educated people being more likely to be aware of the benefits and importance of using contraception. Generally, education is a major determinant of the utilization of health services. This is achieved by improving health literacy and empowering the individual to make informed choices about modern health facilities. Besides, contraceptive use has been shown to affect the timing for fertility among career women. For instance, late marriage due to time spent studying has been documented to facilitate more use of contraception among educated women than illiterates (ibid).

Wealth index was also significantly associated with modern contraceptive use among these married women. Women from the richest tier of the wealth index also had odds that progressively increased to using modern contraceptives among Zambian women. Similar findings regarding the strong association between wealth index and modern contraceptive use were obtained by studies conducted in Malawi and Ghana; women classified within the richest wealth index were more likely to use contraception than those grouped in the poorest index. Financial factors also played an important role in the decreased use of contraceptives among the poorest women. Providing free access for modern contraceptives to poor women would be beneficial to increase their use.^18^

The findings of this study show that slightly more than half of the women aged 15 to 49 (52 percent) used injectables, and the least used male sterilization and other modern methods at 0.1 percent, respectively. Similarly, the study conducted in Tanzania revealed that the most preferred method of contraception in this community was injectables.^19^ A study in Afghanistan showed that religion played a major role in method use and choice.^20^ Islam was strongly correlated with the probability of choosing an Injectable type of contraceptive compared to other modern methods.

## CONCLUSION AND RECOMMENDATIONS

In conclusion, factors such as women’s age, marital status, the number of living children (parity), religion education level, and wealth index remain significant issues in determining modern contraceptive use among women aged 15 to 49 in Zambia. Injectables were the most preferred type of modern contraceptives used among Zambian women aged 15 - 49. Most women intended not to use modern contraceptives any time soon. Female condoms were the most known modern contraceptive.

The study results suggest that policymakers should target certain women and create a campaign regarding modern contraceptive use. Targeting older, poor, and uneducated or less educated women may have a positive impact in terms of increasing their use of modern contraceptives. The intervention should/can take the form of traditional oral or written communication or may involve the use of mobile phones or other technology. FP services may provide counseling as well as improve access to modern contraceptives. Government and the ministry of health should also sensitize the community about different types of modern contraceptives.

## Data Availability

Dataset are available to third parties upon request to Corresponding Author

## AUTHOR CONTRIBUTIONS

CJM conceptualized the work, data acquisition and analysis. TC and MM supervised the analysis and interpretation. CJM, TC, MM and ZM contributed critically to the revision of the manuscript and approval of the final version and are accountable to all aspects of the work regarding its accuracy and integrity.

## Institution Review Board STATEMENT

Ethical clearance was obtained from the University of Lusaka, School of Medicine & Health Sciences Research Ethics Committee Reference: IORG0010092/ MPH1622955. National Health Research Authority (NHRA) granted permission (Ref No. NRA 000016/26/01/2021) in line with the Act of Parliament (number 2 of 2013) which mandates all researchers to submit their research protocols to the NHRA upon receipt of approval from the Research Ethics Committee or an Institutional Review Board. No formal ethical clearance was obtained from ZSA as the study analysed publically available data.

## INFORMED CONSENT STATEMENT

Not applicable

## ACKNOWLEDGEMENTS

This article is part of the Master’s thesis submitted to the University of Lusaka in partial fulfilment of the requirements for the degree of Master of Public Health.

